# Hyperorality in Frontotemporal Dementia: How Psychiatric and Neural Correlates Change Across the Disease Course

**DOI:** 10.1101/2024.02.19.24302699

**Authors:** Christopher B. Morrow, Chiadi Onyike, Alexander Pantelyat, Gwenn S. Smith, Jeannie Leoutsakos, Andreia V. Faria, Neill R. Graff-Radford, R. Ryan Darby, Nupur Ghoshal, Adam M. Staffaroni, Katya Rascovsky, Toji Miyagawa, Akshata Balaji, Kyrana Tsapkini, Maria I. Lapid, Mario F. Mendez, Irene Litvan, Belen Pascual, Julio C. Rojas, Zbigniew K. Wszolek, Kimiko Domoto-Reilly, John Kornak, Vidyulata Kamath, ALLFTD Consortium

**Affiliations:** Department of Psychiatry and Behavioral Sciences, Johns Hopkins University School of Medicine, Baltimore, MD; Department of Neurology, Johns Hopkins University School of Medicine, Baltimore, MD; The Russell Morgan Department of Radiology, Johns Hopkins School of Medicine, Baltimore, MD; Department of Neurology, Mayo Clinic, Jacksonville, FL; Department of Neurology, Vanderbilt University Medical Center, Nashville, TN; Department of Neurology, Washington University School of Medicine, St. Louis, MI; Department of Neurology, Memory and Aging Center, Weill Institute for Neurosciences, University of California, San Francisco, San Francisco, CA; Department of Neurology and Penn Frontotemporal Degeneration Center University of Pennsylvania Perelman School of Medicine, Philadelphia, PA; Department of Neurology, Mayo Clinic, Rochester, MN; Department of Neurology, David Geffen School of Medicine at UCLA, Los Angeles, CA; Department of Neurosciences, UC San Diego, La Jolla, CA; Department of Neurology, Houston Methodist Research Institute, Houston, TX; Department of Neurology, University of Washington School of Medicine, Seattle, WA; Department of Epidemiology and Biostatistics, University of California, San Francisco, San Francisco, CA

**Author notes:** **<u>Correspondence to:</u>**Christopher Morrow, MD, Department of Psychiatry and Behavioral Sciences, The Johns Hopkins University School of Medicine, 600 N. Wolfe Street, Meyer 235, Baltimore, MD 21287.

**Keywords:** Frontotemporal Dementia, Hyperphagia, Psychiatric Symptoms, Neuroimaging

## Abstract

**Objectives:** Hyperorality is one of the core features of behavioral variant frontotemporal dementia (bvFTD), however, the cognitive, psychiatric, and neuroanatomic correlates of hyperorality across disease stages remain unclear. This study works to fill this knowledge gap by exploring these associations in the early and advanced stages of bvFTD.

**Methods:** Participants with sporadic and genetic bvFTD were enrolled in the ARTFL LEFFTDS Longitudinal Frontotemporal Lobar Degeneration consortium (ALLFTD). The primary analyses used baseline data to compare cognitive and psychiatric symptoms in those with and without hyperorality stratified by disease severity. Linear multivariable regressions adjusting for age and total intracranial volume were used to examine baseline associations between regional gray matter volumes and hyperorality status. Five anatomic regions of interest were pre-selected for analysis based on previously identified neuroanatomic correlates of hyperorality in bvFTD.

**Results:** Hyperorality was present in 50% of early-stage bvFTD participants (n = 136) and was associated with higher rates of ritualistic/compulsive behavior. Hyperorality was present in 63% of advanced-stage participants (n = 208) and was associated higher rates of apathy, and ritualistic/compulsive behavior. Regional gray matter volumes were similar in those with and without hyperorality in early-stage participants. In the advanced-stage participants, hyperorality was associated with lower gray matter volumes in the right dorsal and ventral striatum.

**Conclusions:** Hyperorality emerges early in bvFTD and is accompanied by deficits in social cognition and complex-ritualistic behavior prior to clinically significant gray matter volume loss. These findings suggest that early identification and treatment of hyperorality could improve neuropsychiatric trajectories in bvFTD.

## Introduction

The behavioral variant of frontotemporal dementia (bvFTD) is a clinical syndrome characterized by progressive impairments in temperament, behavior, and cognition.[1–3] One of the core features of bvFTD is changes in feeding behavior, often termed hyperorality, which can distinguish FTD from other neurodegenerative conditions.[4] Hyperorality presents as varied combinations of hyperorexia (compulsive overeating), foraging (continuously seeking out food), food fads (restricitive or idiosyncratic food choices), feeding rituals, preference for sweets, food hoarding, alcohol or tobacco binging, and pica.[5] It occurs in approximately 50-60% of individuals with bvFTD, with its occurrence increasing with disease severity, affecting about 50% of individuals with early-stage illness and 75% with advanced illness.[5–8]

The etiology of hyperorality in bvFTD is complex, and various mechanisms have been proposed, including disruptions in frontoinsular, striatal, and orbitofrontal neural networks as well as hypothalamic dysfunction, neuro-endocrine changes, and compensation for a hyper-metabolic state.[1] It has been hypothesized that disruptions in these circuits may lead to dopaminergic dysfunction, especially within the mesocorticolimbic pathways, where similar changes have been linked to impulsivity in other neurodegenerative disesases like Parkinson’s disease, where impulsive overeating can also occur.[9] Serotonergic dysfunction resulting from disruptions in frontoinsular, striatal, and orbitofrontal neural networks has also been linked to impulsivity and may play a role in mediating hyperorality in bvFTD. Atrophy in several neuroanatomic regions has been associated with hyperorality, including the right insula, right striatum, and bilateral orbitofrontal cortices.[10–12] How hyperorality emerges in early-stage bvFTD, however, has not been a focus of prior studies .[10–15] This knowledge gap is of clinical concern and practical consequence, as learning how hyperorality emerges in bvFTD, how individuals with hyperorality differ from those without hyperorality, whether hyperorality represents a prodromal symptom in some cases, and how hyperorality impacts caregiver burden, is key to defining mechanisms and designing targeted therapies.

Given this background, our primary aim was to describe and compare the cognitive, psychiatric, and neuroanatomic profiles of individuals with bvFTD and hyperorality in early and advanced stages of the disease. We tested the hypothesis that individuals with hyperorality will show more varied and severe psychiatric symptom profiles compared to those without hyperorality; specifically, higher levels of compulsive/ritualistic behavior, anxiety, psychosis, and elation, but similar cognitive profiles to those without hyperorality in both early and advanced disease states.[12] The secondary aim was to identify neuroanatomic correlates of hyperorality in bvFTD across stages of the disease using structural magnetic resonance imaging (MRI). We tested the hypothesis that individuals with hyperorality will show atrophy in the following regions of interest: right orbitofrontal cortex, right insula, and right dorsal and ventral striatum, and that atrophy differences would be more pronounced in advanced-stage disease than in early-stage disease.

## Methods

### Participants

Participants were enrolled in ARTFL LEFFTDS Longitudinal Frontotemporal Lobar Degeneration consortium (ALLFTD ) – a multisite study that includes participants from the Advancing Research and Treatment for Frontotemporal Lobar Degeneration (ARTFL) and Longitudinal Evaluation of Familial Frontotemporal Dementia Subjects (LEFFTDS) studies. Each participant underwent extensive clinical interviews and examinations, structural MRI, and blood biomarker sampling. The study protocol and procedures can be found in earlier papers.[16, 17] Participants with a primary clinical diagnosis of bvFTD were included in the analyses, and all those with a primary diagnosis other than bvFTD or anatomical abnormalities on the MRI scan (e.g. tumor or stroke) were excluded. Disease severity was defined based on Clinical Dementia Rating (CDR®) plus NACC FTLD Behavior & Language Domains global score (CDR® plus NACC FTLD).[18, 19] Participants with CDR® plus NACC FTLD scores of ≤1 were classified as early-stage and those with CDR® plus NACC FTLD scores of 2 or 3 as advanced-stage. A separate sensitivity analysis of participants with CDR® plus NACC FTLD equal to 0.5 was also performed.

### Clinical Assessment

The hyperorality variable was drawn from Uniform Data Set (UDS) Form B9F – Clinical PPA and bvFTD Features, part of the FTLD Module.[20] This module was implemented by the NIA Alzheimer’s Disease Research Centers Program (ADRC) to help differentiate the neuropsychological characteristics of FTD from Alzheimer’s disease (AD). The FTLD-CDR module includes a series of psychometric tests sensitive to the behavioral and language impairments common in FTD syndromes (i.e., bvFTD and primary progressive aphasia) and is described in detail in earlier studies.[21] Hyperorality was recorded as present if the hyperorality variable was marked as “definitely present” and absent otherwise. There were 57 cases where hyperorality was marked as “questionable” and these were analyzed as absent hyperorality to avoid including uncertain cases. Sensitivity analyses were performed to include those marked as “questionable” in the hyperorality category, however the overall results did not change meaningfully. Cognitive data were derived from the UDS Version 3.0 neuropsychological battery.[22] Cognitive performance was assessed using standard tests of processing speed (Trail Making Test Part A and Digit Symbol Substitution Test), language (noun naming, category fluency), executive functioning (Trail Making Test B minus Trail Making Test A, Digit Span Backward), memory (Craft Story - Immediate and Delayed) and attention (Digit Span Forward). See supplementary materials for a description of each behavioral scale and cognitive test.

Behavioral scales included measures of self-monitoring, empathetic concern, perspective taking, behavioral inhibition, social norms, depression severity, behavioral functioning, and social behaviors. Data from the Neuropsychiatric Inventory Questionnaire (NPI-Q) captured the following neuropsychiatric symptoms: depression, anxiety, psychosis, apathy, disinhibition, irritability, elation, motor disturbance, and sleep disturbance/nighttime behaviors. The NPI-Q is a validated and widely used questionnaire to assess neuropsychiatric symptoms (NPS) in dementia syndromes, including in bvFTD.[23–27] Neuropsychiatric symptoms were analyzed as dichotomous variables based on their presence or absence on the NPI-Q. We also examined associations between NPS severity and hyperorality. Data on ritualistic/compulsive behavior were drawn from UDS Form B9F – Clinical PPA and bvFTD Features. Ritualistic/compulsive behavior was recorded as present where the variable was marked as “definitely present” and absent otherwise. There were 55 cases recorded as “questionable” and these were analyzed as absent ritualistic/compulsive behavior to avoid including uncertain cases. We examined caregiver burden using the Zarit Burden Interview, a 22-item instrument that is well validated for capturing caregiver burden in FTD spectrum disorders.[28]

### MRI acquisition and Analysis

At all centers, participants underwent T1-weighted structural MRI scans at 3 Tesla using harmonized sequences across multiple vendors (GE, Philips, Siemens) with a standardized imaging protocol similar to that used by the Alzheimer’s Disease Neuroimaging Initiative – versions 2 and 3 (ADNI-2 and ADNI-3 basic) (see https://adni.loni.usc.edu/methods/documents/mri-protocols/).[16] Before image preprocessing, all T1-weighted images underwent quality control (QC) assessment at the Mayo Clinic to ensure that the images were acquired following parameters set by the scanner-specifc MRI protocol and that there were no motion, or other image artifacts that would preclude quantitative analysis. [29] Images that failed QC at any of these steps were excluded from further analysis. T1-weighted images underwent bias field correction using the N3 algorithm, and the segmentation was performed using SPM12 unified segmentation (Wellcome Trust Center for Neuroimaging, London, UK, (http://www.fil.ion.ucl.ac.uk/spm).[30, 31] A customized group template was generated from the segmented gray and white matter tissues and cerebrospinal fluid by non-linear registration template generation using the Large Deformation Diffeomorphic Metric Mapping framework.[32] Native subject space gray and white matter were geometrically normalized to the group template, modulated, and smoothed in the group template. The applied smoothing used a Gaussian kernel with an 8 mm full-width half maximum. Every transformation step was carefully inspected, from the native space to the group template. Baseline cortical volumes were measured for each participant using the Desikan-Killiany cortical atlas.[33]

Five anatomic regions of interest (ROIs) were selected *a priori* based on previously identified neuroanatomic correlates of hyperorality in studies using voxel-based morphometry with a similar population of participants with bvFTD.[11, 12, 15] ROIs included: (1) right lateral orbitofrontal cortex, (2) right medial orbitofrontal cortex, (3) right insula cortex, (4) right dorsal striatum, and (5) right ventral striatum. While other hyperorality-related ROIs have been identified, we focused the analyses on the regions most consistently correlated with feeding behavior in prior FTD studies. As behavioral manifestations, particularly changes in eating behavior, are more often associated with right lateralized atrophy, we restricted our primary analysis to right-sided structures.[12, 34, 35] In secondary exploratory analysis, we also examined additional regions of interest including the left lateral orbitofrontal cortex, left medial oribitofrontal cortex, left insula, left dorsal striatum, left ventral striatum, left and right anterior cingulate, left and right posterior cingulate, left and right frontal lobes, and left and right temporal lobes.

### Statistical Methods

The analyses were based on first-visit data. Differences in participant characteristics and clinical outcomes were compared across groups with and without hyperorality using two-sided t-tests for continuous variables and Pearson χ2 tests for categorical variables. Linear multivariable regression adjusting for age and total intracranial volume (TIV) was used to examine associations between regional gray matter volumes and hyperorality status. We hypothesized that the inclusion of age and total intracranial volume would adequately adjust for individual level variability in brain volumes, however, we also conducted a sensitivity analysis with adjustment for both sex and genetic status in the regression models. Sex and genetic status were generally not associated with significant volumentric differences and as the general trends of our findings were still consistent, the final model did not include adjustment for sex and genetic status in order to avoid over-saturation of the model without meaningfully improving the interpretation of the association of hyperorality on region specific volumentric changes. The primary analyses compared groups with and without hyperorality in both early and advanced-stage disease severity as disease severity was hypothesized to modify the effect of hyperorality on regional gray matter volumes. The statistical significance level, Alpha, was set at 0.05. We also provide indication of where statistically significant results survive Holm-Bonferroni multiple comparison correction. STATA SE 17 (StataCorp LP, College Station, TX) was used for all analyses.

## Results

### Demographics

Demographic data are shown in Table 1. Of 1,316 participants with baseline data, 354 participants had a primary clinical diagnosis of bvFTD, 344 of whom had data on hyperorality and were included in the analysis. There were 136 participants classified as early-stage, with the remaining 208 classified as advanced-stage. Hyperorality was more common in the advanced-stage participants than the early-stage participants (63% versus 50%, p = 0.02). The mean age, years of education, and distribution of men versus women was similar between those with and without hyperorality, across both the early and advanced-stage groups. Those with sporadic bvFTD were more likely to exhibit hyperorality compared to those with a specific genetic mutation (c9orf72, GRN, MAPT) in both the early and advanced-stage groups, however, 22% of the population was missing genetic information, making it challenging to draw definitive conclusions about the association of gene status and hyperorality. A sensitivity analysis classifying early-stage as those with a CDR® plus NACC FTLD score of 0.5 did not lead to materially different results.

**Table 1:** Demographic Characteristics and Mutation Status for bvFTD Patients With (HYP+) and Without Hyperorality (HYP-)

|  |  | HYP-<br>*n* = 68 | HYP+<br>*n* = 68 | *p*-value** |
| --- | --- | --- | --- | --- |
|  |  | mean (SD) | mean (SD) |  |
| CDR FTLD ≤1<br>(*n* = 136, 50% Hyp+) | Age (years) | 62.1 (7.5) | 62.7 (7.5) | 0.61 |
|  | Sex (% Men) | 58.8 | 64.7 | 0.50 |
|  | Education level (years) | 16.1 (2.8) | 16.0 (2.4) | 0.76 |
|  | MoCA | 21.1 (5.4) | 22.8 (9.6) | 0.22 |
|  | CDR-SOB Score (mean) | 3.4 (1.5) | 4.0 (1.4) | 0.02 |
|  | *C9orf72* (*n*, %) | 6 (11.8) | 3 (6.4) | 0.36 |
|  | *GRN* (*n*, %) | 7 (13.7) | 3 (6.4) | 0.23 |
|  | *MAPT* (*n*, %) | 9 (17.7) | 3 (6.4) | 0.09 |
|  | Sporadic (*n*, %) | 29 (56.9) | 38 (80.9) | 0.01 |
|  |  | HYP-<br>*n* = 67 | HYP+<br>*n* = 141 | *p*-value** |
|  |  | mean (SD) | mean (SD) |  |
| CDR FTLD >1<br>(*n* = 208, 63% Hyp+) | Age (years) | 61.2 (10.1) | 62.2 (10.1) | 0.50 |
|  | Sex (% Men) | 57.1 | 50.4 | 0.35 |
|  | Education level (years) | 15.4 (2.3) | 15.7 (2.8) | 0.47 |
|  | MoCA | 18.2 (13.5) | 18.9 (18.7) | 0.76 |
|  | CDR-SOB Score (mean) | 8.0 (3.4) | 9.8 (3.5) | **<0.001** |
|  | *C9orf72* (*n*, %) | 19 (32.2) | 19 (17.3) | 0.03 |
|  | *GRN* (*n*, %) | 3 (5.1) | 7 (6.4) | 0.74 |
|  | *MAPT* (*n*, %) | 8 (13.6) | 7 (6.4) | 0.12 |
|  | Sporadic (*n*, %) | 29 (49.2) | 77 (70.0) | 0.008 |

\*t-values for continuous variables, $\chi^2$ for binary variables, adjusted residuals for categorical variables
\*\*p-values that maintained significance after Holm-Bonferroni correction are bolded

### Neuropsychological Variables

Tables 2 and 3 display scores on cognitive and social behavioral measures. Early-stage and advanced-stage participants with hyperorality performed similarly in processing speed, language, executive functioning, memory, and attention compared to participants without hyperorality at a similar disease stage. Early-stage participants with hyperorality had poorer self-monitoring than those without hyperorality (27.6 versus 21.2, p = 0.001). Advanced-stage participants with hyperorality had higher scores on the Social Behavior Observer Checklist than those without hyperorality. supplementary tables 1 and 2 display cognitive and social behavioral measure scores by disease stage in those with and without hyperorality. As expected, those with advanced illness generally had poorer performance across cognitive and behavioral domains regardless of hyperorality status.

**Table 2:**
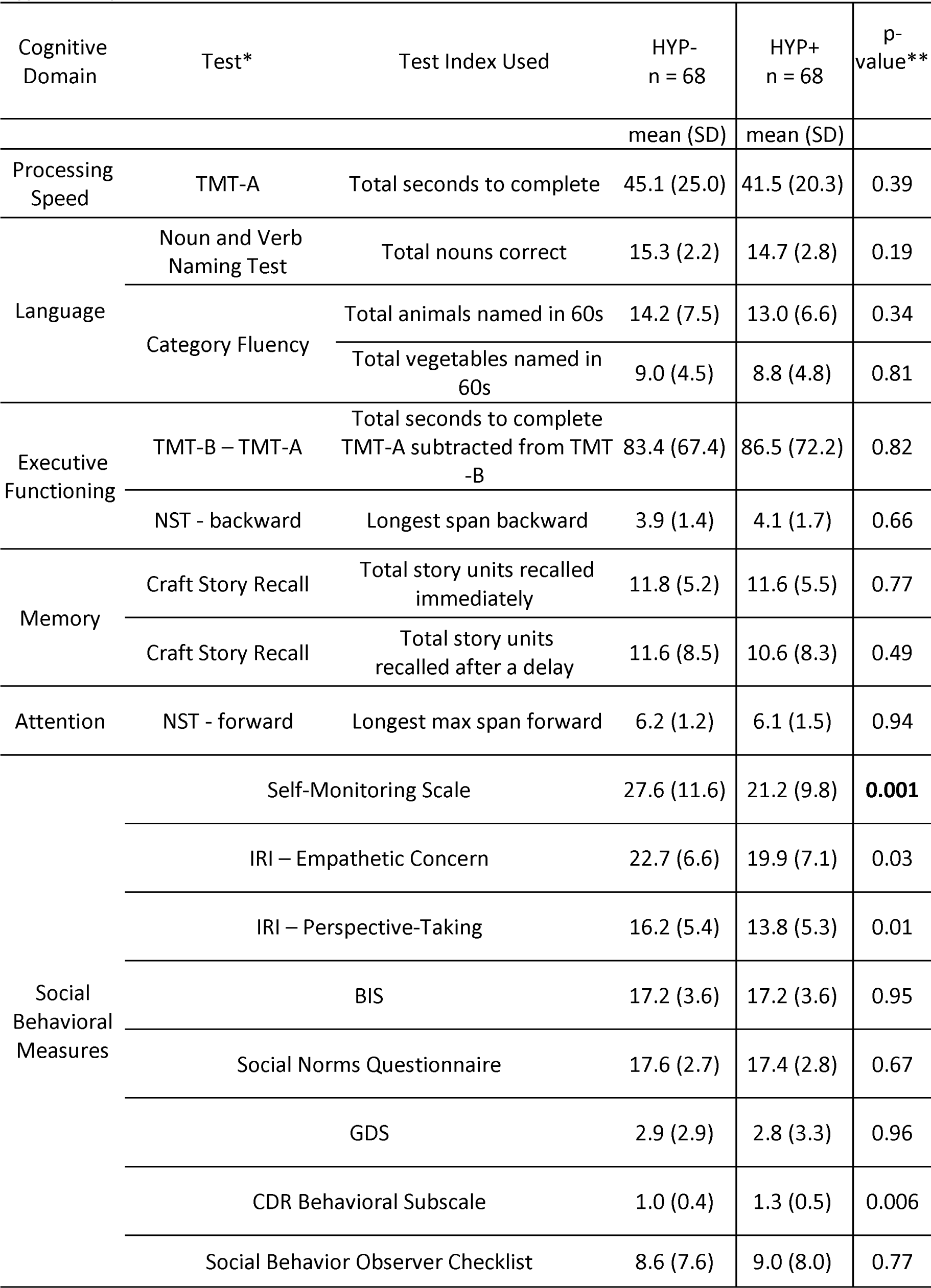
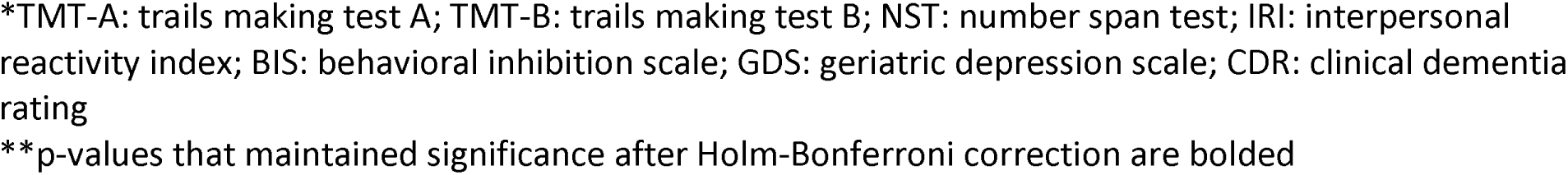
Cognitive and Social Behavioral Scores for Early-Stage bvFTD Patients With (HYP+) and Without Hyperorality (HYP-)

**Table 3:**
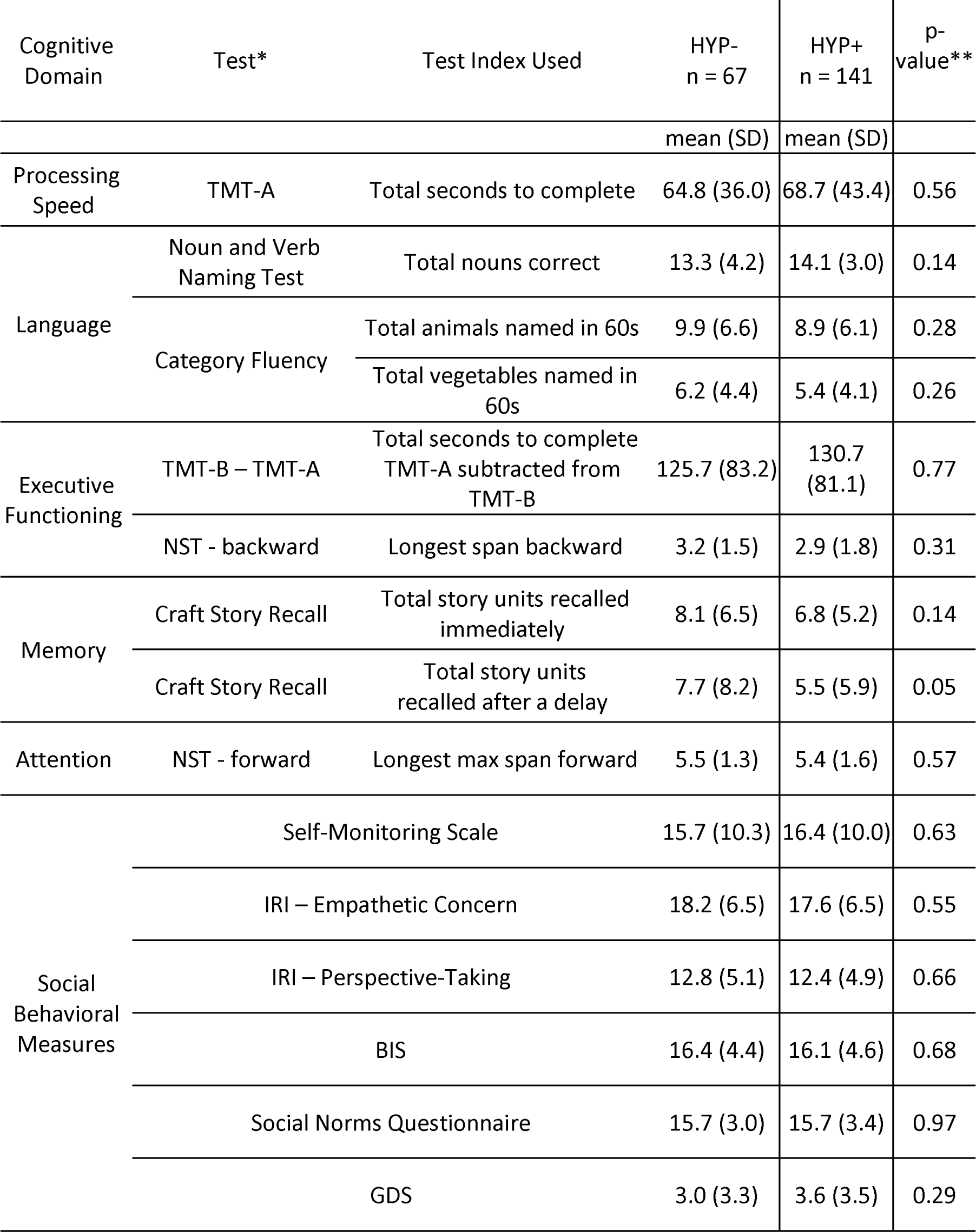
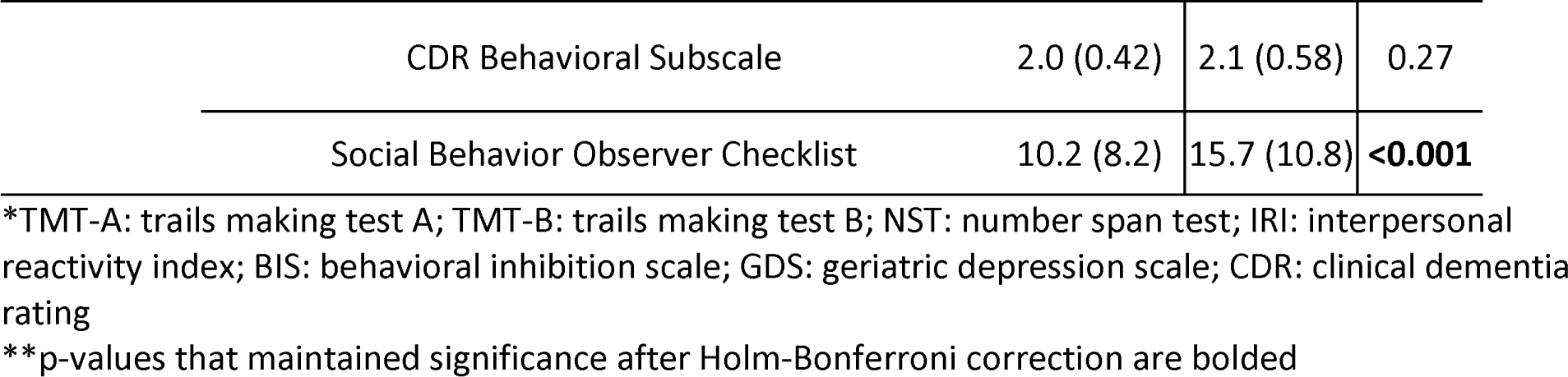
Cognitive and Social Behavioral Scores for Advanced Stage bvFTD Patients With (HYP+) and Without Hyperorality (HYP-)

| Cognitive Domain | Test* | Test Index Used | HYP-<br>n = 67 | HYP+<br>n = 141 | p-value** |
| --- | --- | --- | --- | --- | --- |
|  |  |  | mean (SD) | mean (SD) |  |
| Processing Speed | TMT-A | Total seconds to complete | 64.8 (36.0) | 68.7 (43.4) | 0.56 |
| Language | Noun and Verb Naming Test | Total nouns correct | 13.3 (4.2) | 14.1 (3.0) | 0.14 |
|  | Category Fluency | Total animals named in 60s | 9.9 (6.6) | 8.9 (6.1) | 0.28 |
|  |  | Total vegetables named in 60s | 6.2 (4.4) | 5.4 (4.1) | 0.26 |
| Executive Functioning | TMT-B – TMT-A | Total seconds to complete TMT-A subtracted from TMT-B | 125.7 (83.2) | 130.7 (81.1) | 0.77 |
|  | NST - backward | Longest span backward | 3.2 (1.5) | 2.9 (1.8) | 0.31 |
| Memory | Craft Story Recall | Total story units recalled immediately | 8.1 (6.5) | 6.8 (5.2) | 0.14 |
|  | Craft Story Recall | Total story units recalled after a delay | 7.7 (8.2) | 5.5 (5.9) | 0.05 |
| Attention | NST - forward | Longest max span forward | 5.5 (1.3) | 5.4 (1.6) | 0.57 |
| Social Behavioral Measures | Self-Monitoring Scale |  | 15.7 (10.3) | 16.4 (10.0) | 0.63 |
|  | IRI – Empathetic Concern |  | 18.2 (6.5) | 17.6 (6.5) | 0.55 |
|  | IRI – Perspective-Taking |  | 12.8 (5.1) | 12.4 (4.9) | 0.66 |
|  | BIS |  | 16.4 (4.4) | 16.1 (4.6) | 0.68 |
|  | Social Norms Questionnaire |  | 15.7 (3.0) | 15.7 (3.4) | 0.97 |
|  | GDS |  | 3.0 (3.3) | 3.6 (3.5) | 0.29 |

|  |  |  |  |
| --- | --- | --- | --- |
| CDR Behavioral Subscale | 2.0 (0.42) | 2.1 (0.58) | 0.27 |
| Social Behavior Observer Checklist | 10.2 (8.2) | 15.7 (10.8) | **<0.001** |

\*TMT-A: trails making test A; TMT-B: trails making test B; NST: number span test; IRI: interpersonal reactivity index; BIS: behavioral inhibition scale; GDS: geriatric depression scale; CDR: clinical dementia rating
\*\*p-values that maintained significance after Holm-Bonferroni correction are bolded

### Neuropsychiatric symptoms

Table 4 displays associations between hyperorality and neuropsychiatric symptoms. Early-stage participants with hyperorality displayed higher rates of motor disturbance and ritualistic/compulsive behavior than those without hyperorality (only ritualistic/compulsive behavior survive Holm-Bonferroni correction, Cramer’s V = 0.41). Advanced-stage participants with hyperorality had higher rates of anxiety, elation, apathy (Cramer’s V = 0.30) and ritualistic/compulsive behavior (Cramer’s V = 0.22) than those without hyperorality (only apathy and ritualistic/compulsive behavior survive Holm-Bonferroni correction). Early-stage participants with hyperorality also had higher total NPI-Q scores than those without hyperorality (Cohen’s D = 0.42). supplementary tables 3 and 4 display neuropsychiatric symptom states by disease stage in participants with and without hyperorality. In participants without hyperorality, those with advanced-stage illness had higher rates of ritualistic/compulsive behavior (Cramer’s V = 0.32) than those with early-stage illness. In those with hyperorality, depression was more common in early-stage participants (Cramer’s V = 0.23). Caregiver burden as captured by the Zarit Burden Interview is displayed in Table 5. Caregivers of patients with early-stage disease and hyperorality scored higher overall on this scale than those without hyperorality, however, there was not evidence of an unequivocal difference (total score 33 versus 38, p-value 0.06). Across specific domains of caregiver burden, there was evidence of increased burden in certain areas when hyperorality was present in early-stage disease. Caregiver’s of early-stage participants with hyperorality were more likely to report that their health had suffered because of their involvement with their loved one (mean score 1.2 versus 1.6, p-value 0.03), that their social life had suffered (mean score 1.5 versus 2.0, p-value 0.03), that they felt uncomfortable hosting friends (mean score 0.9 versus 1.4, p-value 0.02), and feeling a loss of control in life (mean score 1.1 versus 1.7, p-value 0.003). Caregiver’s of late-stage participants with hyperorality were more likely to report that they do not have enough time for themselves (mean score 2.0 versus 2.5, p-value 0.008), and that they feel stressed about meeting other responsibilities related to family or work ( mean score 2.4 versus 2.8, p-value 0.03).

**Table 4:** Association of Neuropsychiatric Symptoms and Hyperorality Across Disease States of bvFTD.

|  |  | HYP- | HYP+ | Difference<br>(95% CI) | p-value* |
| --- | --- | --- | --- | --- | --- |
|  |  | % | % |  |  |
| CDR FTLD ≤1 | Psychosis | 18.2 | 25.4 | 7.2<br>(-7.0 – 21.4) | 0.32 |
|  | Agitation | 43.3 | 56.9 | 13.6<br>(-3.2 – 30.5) | 0.12 |
|  | Depression | 51.5 | 47.7 | -3.8<br>(-20.9 – 13.3) | 0.66 |
|  | Anxiety | 48.5 | 51.6 | 3.1<br>(-14.1 – 20.3) | 0.73 |
|  | Elation | 22.7 | 35.4 | 12.7<br>(-2.7 – 28.1) | 0.11 |
|  | Apathy | 77.3 | 73.9 | 3.4<br>(-18.1 – 11.3) | 0.65 |
|  | Disinhibition | 56.1 | 73.9 | 14.8<br>(-1.2 – 30.7) | 0.07 |
|  | Irritability | 61.5 | 66.2 | 4.6<br>(-11.9 – 21.1) | 0.58 |
|  | Motor Disturbance | 42.4 | 64.6 | 22.2<br>(5.5 – 38.8) | 0.01 |
|  | Sleep Disturbance | 41.7 | 51.6 | 9.9<br>(-7.7 – 27.6) | 0.27 |
|  | Ritualistic/Compulsive Behavior | 25.0 | 66.2 | **41.2**<br>**(25.9 – 56.4)** | **<0.001** |
|  | NPI-Q Total score [mean (SD)] | 9.3 (5.9) | 11.8 (6.4) | **2.6**<br>**(0.30 – 4.8)** | **0.03** |
| CDR FTLD >1 | Psychosis | 23.3 | 23.8 | 0.5<br>(-11.7 – 12.7) | 0.93 |
|  | Agitation | 56.8 | 52.8 | 4.0<br>(-18.2 – 10.2) | 0.58 |
|  | Depression | 36.5 | 24.8 | -11.7<br>(-25.0 – 1.6) | 0.08 |
|  | Anxiety | 35.1 | 55.2 | 20.0<br>(6.1 – 34.0) | 0.006 |
|  | Elation | 13.5 | 31.0 | 17.4<br>(6.2 – 28.7) | 0.006 |
|  | Apathy | 64.0 | 88.9 | **24.9**<br>**(12.7 – 37.1)** | **<0.001** |
|  | Disinhibition | 67.6 | 75.0 | 7.4<br>(-5.7 – 20.5) | 0.26 |
|  | Irritability | 56.0 | 56.7 | 0.7<br>(-13.5 – 14.9) | 0.92 |
|  | Motor Disturbance | 62.7 | 69.3 | 6.6<br>(-6.9 – 20.2) | 0.33 |
|  | Sleep Disturbance | 46.0 | 50.8 | 4.8<br>(-9.5 – 19.2) | 0.51 |
|  | Ritualistic/Compulsive Behavior | 57.1 | 77.9 | **20.7**<br>**(7.6 – 33.9)** | **0.002** |
|  | NPI-Q Total score [mean (SD)] | 10.4 (7.3) | 12.6 (6.9) | 2.1<br>(0.03 – 4.3) | 0.05 |

\*p-values that maintained significance after Holm-Bonferroni correction are bolded

**Table 5:**
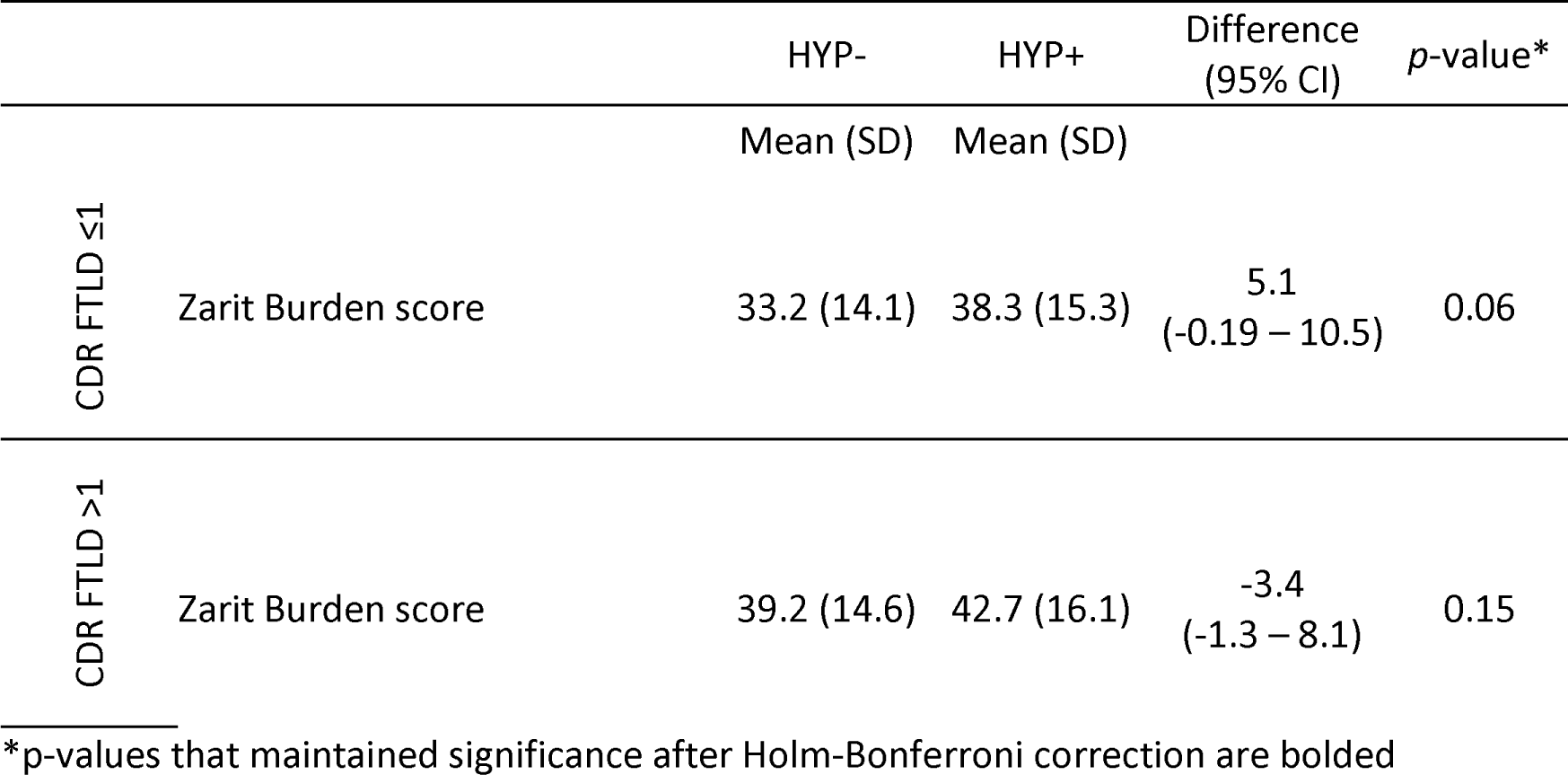
Neuropsychiatric Symptom Severity and Hyperorality Across Disease States in bvFTD.

|  |  | HYP- | HYP+ | Difference<br>(95% CI) | p-value* |
| --- | --- | --- | --- | --- | --- |
|  |  | Mean (SD) | Mean (SD) |  |  |
| CDR FTLD ≤1 | Zarit Burden score | 33.2 (14.1) | 38.3 (15.3) | 5.1<br>(-0.19 – 10.5) | 0.06 |
| CDR FTLD >1 | Zarit Burden score | 39.2 (14.6) | 42.7 (16.1) | -3.4<br>(-1.3 – 8.1) | 0.15 |

\*p-values that maintained significance after Holm-Bonferroni correction are bolded

### Gray Matter Regions of Interest (ROI)

MRI-based volumetric measurements of relevant brain structures were available for 98 participants with hyperorality data and were used in the analysis of first visit data. Demographic characteristics of those with MRI and hyperorality data are displayed in supplementary table 7. The sub-sample with MRI data appears to be largely representative of the study sample. Of those with MRI data, advanced-stage participants without hyperorality tended to be younger than advanced-stage participants with hyperorality, however, we adjusted for age in our analyses of ROI gray matter volume to account for age differences. Associations between hyperorality and gray matter volumes are displayed in Table 6. Contrary to expectation, regional gray matter volumes were similar in those with and without hyperorality in early-stage participants. In the advanced-stage participants, hyperorality was associated with lower gray matter volumes in the right dorsal striatum (hyperorality coefficient -692.6, 95% CI [-1194.3 – -190.9], p-value 0.008, eta-squared 0.14), and ventral striatum (hyperorality coefficient -36.0, 95% CI [-62.8 – -9.2], p-value 0.009, eta-squared 0.13). This effect was attenuated when those with “questionable” hyperorality were included in the hyperorality group (see supplementary table 10), however, given the goal of studying those with clear hyperorality symptoms, the final analysis included only those with definitive hyperorality. The effect of hyperorality on regional gray matter volume was also attenuated when adjustments for sex and genetic status were included in the regression model, however, given that sex and genetic status were not significantly associated with volumetric differences, and that age and total intracranial volume appear to adjust appropriately for individual volumetric variability, a final model without sex and genetic status was used. Similarly, we found no association between handedness and hyperorality or handedness and volumetric changes in regions of interest, so did not include handedness in the final regression model. supplementary tables 8 and 9 display associations between disease stage and gray matter volumes in those with and without hyperorality to account for volumetric changes related to the disease stage. Regional gray matter volumes were similar in participants with and without hyperorality based on disease stage. No exploratory regions of interest showed definitive volumetric differences based on the presence of hyperorality.

**Table 6:**
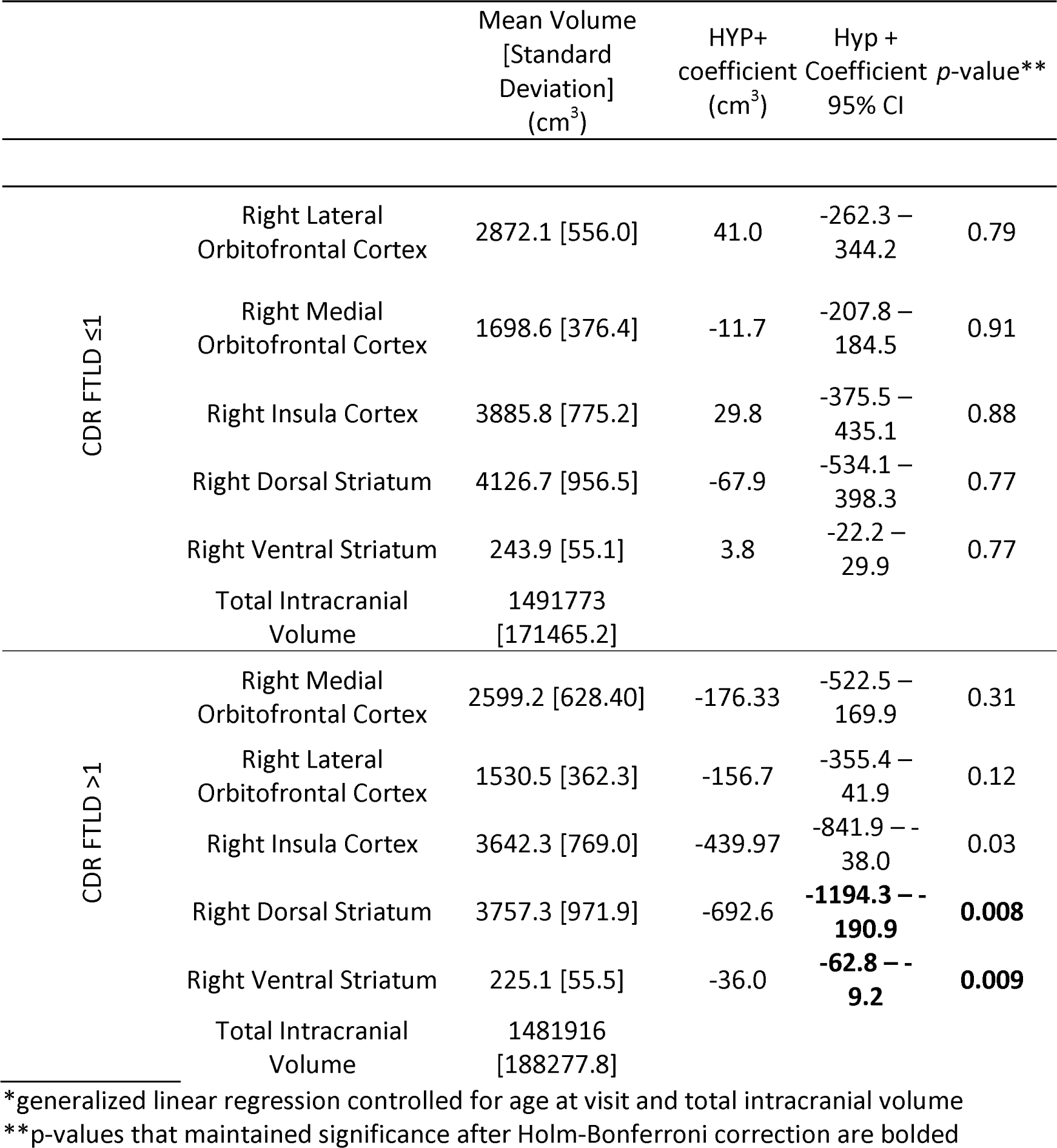
Neuroanatomic Changes and Hyperorality in Early and Advanced bvFTD for Primary Regions of Interest.

## Discussion

### Overview

Our study examined the cognitive, psychiatric, and neuroanatomic profiles of participants with bvFTD and hyperorality at early and advanced stages of illness. As hypothesized, we found that individuals with hyperorality had more varied and severe psychiatric symptom profiles than those without hyperorality, and that psychiatric symptom profiles in those with hyperorality differ across the disease course. Also, as hypothesized, we found similar cognitive profiles in those with and without hyperorality in both early and advanced disease states across all cognitive domains tested (processing speed, language, executive functioning, memory, and attention). Contrary to expectations, we did not identify unequivocal neuroanatomic changes in pre-specified regions of interest in those with hyperorality and early-stage disease compared to those without hyperorality. However, there was more atrophy in advanced-stage participants with hyperorality compared to those without hyperorality in the right dorsal and ventral striatum.

### Psychiatric Symptoms

In early-stage participants, we found that hyperorality was associated with higher rates of motor disturbances and ritualistic/compulsive behaviors compared to those without hyperorality. In advanced-stage participants with hyperorality we observed higher rates of anxiety, elation, ritualistic/compulsive behavior and apathy relative to those without hyperorality. Prior work has observed similar behaviors in obsessive-compulsive disorder (OCD) in connection with dysfunctional cortico-striatal circuits.[36] Elevated rates of obsessive-compulsive disorder in individuals with eating disorders have also been observed, supporting the possibility of a shared underlying neurobiology between hyperorality and OCD.[37, 38]

### Cognitive Profiles

Cognitive profiles were comparable in those with and without hyperorality in both early and advanced disease states. This finding is consistent with observations from smaller studies and aligns with the view that hyperorality is not a by-product of cognitive impairment but rather a behavioral phenotype arising from a particular pattern of neurodegeneration.[7, 12] We did observe a reduced ability to detect and react to social and emotional expressions in the early-stage participants with hyperorality compared to those without hyperorality. In addition, we observed an increase in aberrant behavior during neuropsychological testing sessions (as evidenced by elevated Social Behavior Observer Checklist scores) in the advanced-stage participants with hyperorality compared to those without hyperorality but not in those with early-stage disease. These observations suggest that social behavior declines earlier in participants with hyperorality than those without hyperorality, and that in advanced illness those with hyperorality exhibit more impaired social behavior in clinical encounters than those with hyperorality in early-stage FTD.

### Role of Disease Severity

The neuropsychiatric symptoms and neuroanatomic changes in participants with hyperorality do not appear to be explained by differences in disease severity alone. To test this possibility, we performed separate analyses comparing neuropsychiatric symptoms and gray matter volumes in regions of interest in participants with and without hyperorality, stratified by disease severity. While those with hyperorality showed more depression in early-stage illness, the elevated rates of ritualistic/compulsive behavior and apathy in the advanced stage group with hyperorality compared to the advanced stage group without hyperorality did not appear to be explained by disease severity. In addition, disease severity did not explain the lower volumes observed in the right dorsal striatum and right ventral striatum in the advanced stage participants with hyperorality compared to the advanced stage participants without hyperorality.

### Implications for Understanding Hyperorality in bvFTD

Taken together, our findings provide three intriguing ideas about the nature and course of aberrant feeding behavior in bvFTD. First, the observation that hyperorality occurs at high rates in early as well as advanced stage bvFTD lends support to the notion that hyperorality is an integral phenomenon in the early bvFTD phenotype, providing support for its place in the formal diagnostic criteria.[3] As approximately half of subjects did not exhibit hyperorality early on, further work is needed to clarify whether subjects with early hyperorality represent a particular behavioral phenotype with a distinct underlying neurobiology and trajectory. Second, the lack of association of hyperorality with loss of gray matter volume in the right dorsal striatum and ventral striatum in early-stage illness suggests that changes in functional connectivity of neural circuits likely precedes structural changes. Future studies are needed to understand the structural changes associated with hyperorality in regions outside of the ROIs evaluated as well as to map changes in neural circuitry associated with early bvFTD to delineate early neural substrates of hyperorality. Once identified, these substrates may be informative for bvFTD, and for eating disorders in other conditions like Prader Willi, binge eating disorder, and other conditions marked by hyperphagia and hyperorality. Third, the observation that early-stage participants with hyperorality diverge behaviorally from those without hyperorality, prior to showing structural neuroanatomic change, suggests that there is a window for intervention prior to significant neurodegeneration.

BvFTD manifestations of hyperorality and a compulsion towards oral behaviors hold diagnostic and therapeutic implications. Its presence not only acts as a hallmark for bvFTD but also offers insight into disease progression. For caregivers and clinicians, understanding this symptom is essential in devising tailored interventions and ensuring patient safety. In the context of complexities of bvFTD symptom progression, there is an urgent need to find effective therapeutic interventions.

### Designing Precision Therapies: Implications for Non-Invasive Brain Stimulation

While hyperorality stands out as a distinct and challenging symptom in FTD, recent advances offer hope in managing it. The association of hyperorality and ritualistic/compulsive behavior, alongside striatal gray matter volume loss, points to a possible target for interventions and one promising avenue that is gaining traction is the application of non-invasive brain stimulation. Non-invasive brain stimulation techniques, including transcranial magnetic stimulation and transcranial direct current stimulation (tDCS) targeting the cortico-striato-thalamo-cortical loops by stimulating areas like the dorsolateral prefrontal cortex (dlPFC), have been used successfully in treating refractory symptoms of obsessive-compulsive disorder.[39, 40] Similarly, non-invasive brain stimulation has been used successfully in eating disorders, including binge eating and obesity, targeting the salience network and cortico-striatal-thalamic network through the dlPFC.[41] Non-invasive brain stimulation paradigms like those used in OCD and binge eating could have potential in treating hyperorality in early-stage bvFTD by targeting the cortico-striato-thalamo-cortical networks through stimulation of the dlPFC.

Invasive brain stimulation techniques including deep brain stimulation (DBS) have also been used for severe refractory cases of OCD, targeting the ventral striatum or nucleus accumbens.[42, 43] Similar targets for deep brain stimulation are being explored for treatment of addiction and pathologic overeating.[44, 45] While our work does not explore the functional neuroanatomic changes underlying hyperorality in early-stage bvFTD, the potential for novel therapies is intriguing.

tDCS paired with speech therapy has been used successfully in FTD language variants (non-fluent/agrammatic primary progressive aphasia), improving language abilities. This approach is a model that could be used in bvFTD to target behavioral symptoms including hyperorality.[46, 47] By targeting relevant neural networks, it may be possible to improve feeding behavior. In bvFTD, hyperorality can pose significant risks to health including ingestion of toxic items. Further work is needed to understand the changes in neural circuitry associated with early-stage bvFTD and to determine whether stimulation of relevant networks can slow behavioral deterioration.

While our study benefits from a large population with well characterized and validated diagnoses of bvFTD, some limitations merit consideration. Aspects of hyperorality were not assessed, which limits assessment of types and severity of abnormal feeding behavior; relying on a binary assessment of hyperorality does not fully characterize the behavior or its severity. Future studies will benefit from use of more sophisticated assessments such as the Appetite and Eating Habits Questionnaire (APEHQ), the Disinhibition, Apathy, Perseverations, Hyperorality, Neglect, Empathy (DAPHNE) scale, or the Cambridge Behavioral Inventory.[5, 48–50] However, the finding that the volumetric differences in those with and without hyperorality are more significant when only those with definitive hyperorality are included in the hyperorality category, suggests that more severe hyperorality symptoms are associated with clearer neuroanatomical changes. Our study population consists predominantly of white men living in North America, which may limit the generalizability of our results to other ethnocultural groups and geographic locations. Our study also focuses on baseline visits and pre-specified regions of interest – future longitudinal studies with unbiased voxel-based morphometry analysis are needed to build on our findings. Finally, as with all multisite imaging studies, results can be biased by the use of selective protocols at different centers, however, this bias was minimized by using standardized imaging protocols with centralized and systematic quality control.

## Conclusion

In this study, we have shown that hyperorality emerges early in bvFTD and is accompanied by behavioral and neuropsychiatric symptoms prior to significant gray matter volume loss. In advanced-stage illness, hyperorality is associated with more varied neuropsychiatric symptoms as well as lower gray matter volumes in striatal brain regions. While there are no current disease modifying therapies in FTD, our findings suggest that there is a window for intervention in early-stage bvFTD. Future studies are needed to better understand the early functional neuroanatomic changes and the potential for their translation into targets for novel interventions like non-invasive brain stimulation.

## Data Availability

Data used in this study are from the ALLFTD consortium. Data requests can be submitted at: https://mayojointresearch.sjc1.qualtrics.com/jfe/form/SV_8eTGP626bMRIbyK

https://www.allftd.org

## Author Contributions

All authors contributed to the design and/or analytic approach of the study. CM performed initial analyses and drafted the manuscript text. All other authors reviewed each draft and made substantive revisions and approved the final manuscript.

## Conflicts of Interest

Preliminary analysis from this work was presented as an abstract at the ACTS Conference in Washington, DC on April 18, 2023. CM is funded by KL2TR003099. AP is funded by NIA/NINDS (K23AG059891 and U01NS102035). JR is funded by NIA/NIH K23AG059888, AlzOut, Shenandoah fund and Jon and Gale Love Alzheimer’s fund and is a site PI for clinical trials sponsored by Eli-Lilly and Eisai and a consultant and speaker for Roon Health, Inc. ZKW is partially supported by the NIH/NIA and NIH/NINDS (1U19AG063911, FAIN: U19AG063911), Mayo Clinic Center for Regenerative Medicine, the gifts from the Donald G. and Jodi P. Heeringa Family, the Haworth Family Professorship in Neurodegenerative Diseases fund, The Albertson Parkinson’s Research Foundation, and PPND Family Foundation. He serves as PI or Co-PI on Biohaven Pharmaceuticals, Inc. (BHV4157-206) and Vigil Neuroscience, Inc. (VGL101-01.002, VGL101-01.201, PET tracer development protocol, Csf1r biomarker and repository project, and ultra-high field MRI in the diagnosis and management of CSF1R-related adult-onset leukoencephalopathy with axonal spheroids and pigmented glia) projects/grants. He serves as Co-PI of the Mayo Clinic APDA Center for Advanced Research and as an external advisory board member for the Vigil Neuroscience, Inc., and as a consultant on neurodegenerative medical research for Eli Lilli & Company. IL’s research is supported by the National Institutes of Health grants: 2R01AG038791-06A, U01NS100610, R25NS098999; U19 AG063911-1 and 1R21NS114764-01A1; the Michael J Fox Foundation, Parkinson Foundation, Lewy Body Association, CurePSP, Roche, Abbvie, Biogen, Lundbeck, EIP-Pharma, Biohaven Pharmaceuticals, Novartis, and United Biopharma SRL, UCB. She is a member of the Scientific Advisory Board for Amydis, but does not receive funds and from the Rossy PSP Program at the University of Toronto. She receives her salary from the University of California San Diego and as Chief Editor of Frontiers in Neurology. VK is partially supported by the NIH/NIA and NIH/NINDS (R01AG064093, R01NS108452).

## Supplementary Material: Cognitive Domains and Social Behavioral Scales

### Processing Speed

- Trails Making Test[51]

◦ This test is used as an indicator of visual scanning, graphomotor speed, and executive function. The test consists of two parts (A and B). In part A, the respondent must connect randomly arranged circles containing numbers from 1 to 25 in the correct sequence. In part B, the task is similar, however, the respondent must alternate between letters and numbers.
- Digit Symbol Substitution Test[52]

◦ Requires respondent to match symbols with their corresponding numbers over a 90 second time interval.

### Language

- Noun and Verb Naming Test[53]

◦ A confrontation naming test that is composed of 60 black and white drawings that are ordered from easy to difficult.
- Category Fluency[54]

`◦` Animals: total number of animals named in 60 seconds
◦ Vegetables: total number of vegetables named in 60 seconds

### Executive Functioning

- Trails Making Part A & B[51] (see above)
- Digit Span Backward[51]

◦ Respondents are asked to repeat series of digits that become gradually longer. The maximum digit span that the respondent can repeat backward constitutes the digit span backward score.

### Memory

- Logical Memory Immediate[55]

◦ Total number of story units recalled immediately after a respondent hears a story.
- Logical Memory Delayed

◦ Total number of story units recalled following a delay after a respondent hears a story.

### Attention

- Digit Span Forward[51]

◦ Respondents are asked to repeat series of digits that become gradually longer. The maximum digit span that the respondent can repeat forward constitutes the digit span forward score.

### Social Behavioral Measures

- Self-Monitoring Scale[56]

◦ 13-item informant questionnaire that measures responsiveness to emotional expressions during interactions.
- Empathic Concern Scale[57]

◦ Derived from the Interpersonal Reactivity Index which measure cognitive and emotional components of empathy. It consists of 28 items with two seven-item subscales measuring cognitive empathy including an empathic concern score.
- Perspective Taking Scale[57]

◦ Derived from the Interpersonal Reactivity Index which measure cognitive and emotional components of empathy. It consists of 28 items with two seven-item subscales measuring cognitive empathy including a perspective taking score.
- Behavioral Inhibition Scale[21]

◦ A scale that measures behavioral inhibition by assessing withdrawal-related behavior traits including self-criticism, introversion, and social anxiety.
- Social Norms Questionnaire[58]

◦ 22-item questionnaire used to detect inappropriate social behavior in prompted scenarios. Goal is to measure the degree to which a subject understands widely accepted social boundaries in American culture.
- Geriatric Depression Scale[59]

◦ A self-rating screening tool to measure depressive symptoms in older adults.
- CDR Behavioral Subscale[60]

◦ A subcale developed along with a language subscale to more accurately reflect the clinical status of patients with non-Alzheimer’s dementia like bvFTD.
- Social Behavior Observer Checklist[21]

◦ A checklist which notes the frequency of spontaneous behaviors during clinical evaluation that the examiner observes – these include odd or inappropriate behavior.

**Supplementary Table 1:**
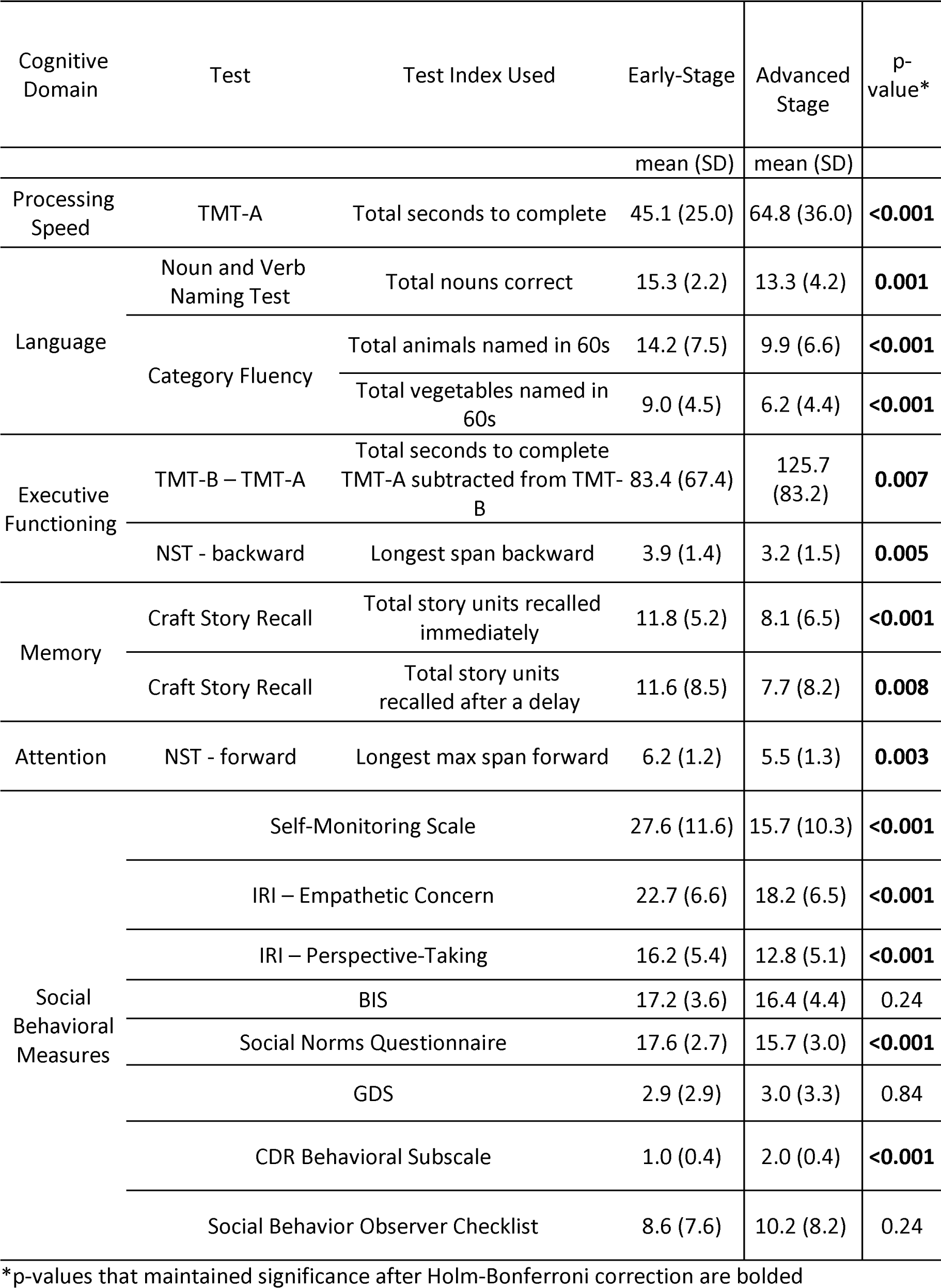
Cognitive and Social Behavioral Scores in Participants without Hyperorality by Disease Stage.

**Supplementary Table 2:**
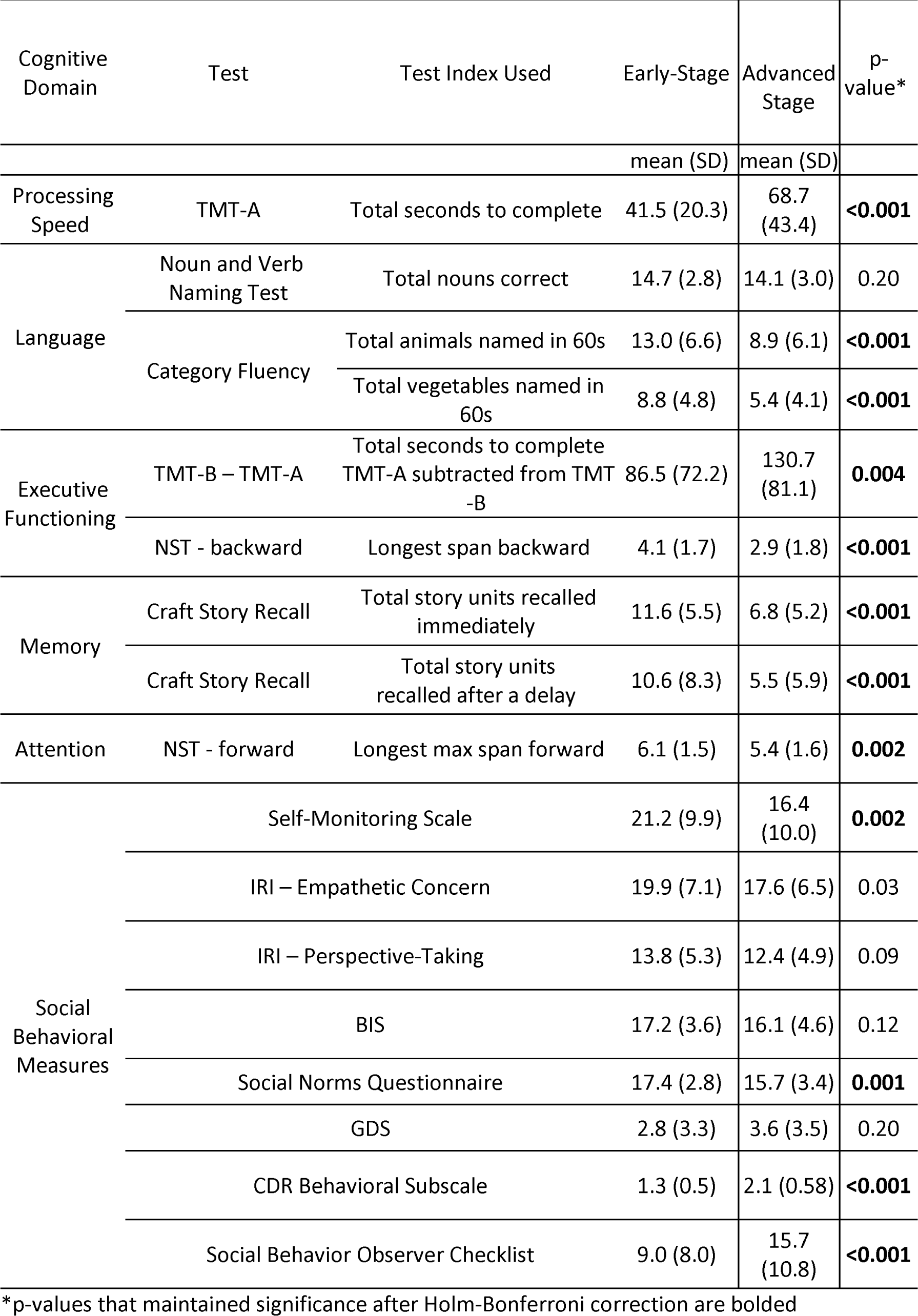
Cognitive and Social Behavioral Scores in Participants with Hyperorality by Disease Stage.

**Supplementary Table 3:** Neuropsychiatric Symptoms in Participants without Hyperorality by disease stage.

|  | Early-Stage | Advanced Stage | *p*-value* |
| --- | --- | --- | --- |
|  | % | % |  |
| Psychosis | 18.2 | 23.3 | 0.46 |
| Agitation | 43.3 | 56.8 | 0.11 |
| Depression | 51.5 | 36.5 | 0.07 |
| Anxiety | 48.5 | 35.1 | 0.11 |
| Elation | 22.7 | 13.5 | 0.16 |
| Apathy | 77.3 | 64.0 | 0.09 |
| Disinhibition | 59.1 | 67.6 | 0.30 |
| Irritability | 61.5 | 56.0 | 0.51 |
| Motor Disturbance | 42.4 | 62.7 | 0.02 |
| Sleep Disturbance | 41.7 | 46.0 | 0.62 |
| Ritualistic/Compulsive Behavior | 25.0 | 57.1 | **<0.001** |

\*p-values that maintained significance after Holm-Bonferroni correction are bolded

**Supplementary Table 4:** Neuropsychiatric Symptoms in Participants with Hyperorality by disease stage.

|  | Early-Stage | Advanced Stage | *p*-value* |
| --- | --- | --- | --- |
|  | % | % |  |
| Psychosis | 25.4 | 23.8 | 0.81 |
| Agitation | 56.9 | 52.8 | 0.58 |
| Depression | 47.7 | 24.8 | **0.001** |
| Anxiety | 51.6 | 55.2 | 0.64 |
| Elation | 35.4 | 31.0 | 0.54 |
| Apathy | 73.9 | 88.9 | 0.008 |
| Disinhibition | 73.9 | 75.0 | 0.86 |
| Irritability | 66.2 | 56.7 | 0.21 |
| Motor Disturbance | 64.6 | 69.3 | 0.51 |
| Sleep Disturbance | 51.6 | 50.8 | 0.92 |
| Ritualistic/Compulsive Behavior | 66.2 | 77.9 | 0.08 |

\**p*-values that maintained significance after Holm-Bonferroni correction are bolded

**Supplementary Table 5:** Neuropsychiatric Symptom Severity in Participants without Hyperorality by disease stage.

|  | Early-Stage | Advanced<br>Stage | *p*-value |
| --- | --- | --- | --- |
|  | Mean (SD) | Mean (SD) |  |
| NPI-Q Total score | 9.3 (5.9) | 10.4 (7.3) | 0.33 |

**Supplementary Table 6:** Neuropsychiatric Symptom Severity in Participants with Hyperorality by disease stage.

|  | Early-Stage | Advanced Stage | *p*-value |
| --- | --- | --- | --- |
|  | Mean (SD) | Mean (SD) |  |
| NPI-Q Total score | 11.8 (6.4) | 12.6 (6.9) | 0.48 |

**Supplementary Table 7:** Demographic Characteristics and Mutation Status for bvFTD Patients With (HYP+) and Without Hyperorality (HYP-) – Subsample with MRI data available.

|  |  | HYP-<br>*n* = 27 | HYP+<br>*n* = 18 | *p*-value** |
| --- | --- | --- | --- | --- |
|  |  | mean (SD) | mean (SD) |  |
| CDR FTLD ≤1 (*n* = 136, 50% Hyp+) | Age (years) | 60.1 (8.0) | 62.2 (7.4) | 0.37 |
|  | Sex (% Men) | 51.9 | 55.6 | 0.81 |
|  | Education level (years) | 15.7 (2.5) | 16.3 (1.7) | 0.33 |
|  | MOCA | 22.1 (4.1) | 22.3 (4.8) | 0.88 |
|  | CDR Global (mean) | 0.6 (0.2) | 0.7 (0.3) | 0.09 |
|  | CDR-FTLD (mean) | 0.8 (0.2) | 1.0 (0.1) | 0.03 |
|  | C9orf72 (% , *n* = 4) | 13.3 | 40.0 |  |
|  | GRN (% , *n* = 9) | 40.0 | 60.0 | 0.14 |
|  | MAPT (% , *n* = 7) | 46.7 | 0.0 |  |
|  |  | HYP-<br>*n* = 19 | HYP+<br>*n* = 34 | *p*-value |
|  |  | mean (SD) | mean (SD) |  |
| CDR FTLD >1 (*n* = 208, 63% Hyp+) | Age (years) | 57.1 (10.2) | 65.1 (7.8) | **0.002** |
|  | Sex (% Men) | 63.2 | 44.1 | 0.40 |
|  | Education level (years) | 14.8 (2.4) | 15.8 (2.6) | 0.21 |
|  | MOCA | 17.1 (6.6) | 17.9 (14.2) | 0.81 |
|  | CDR Global (mean) | 1.1 (0.37) | 1.4 (0.70) | 0.04 |
|  | CDR-FTLD (mean) | 2.0 (0) | 2.1 (0.29) | 0.19 |
|  | C9orf72 (% , *n* = 11) | 61.5 | 30.0 |  |
|  | GRN (% , *n* = 2) | 0 | 20.0 | 0.14 |
|  | MAPT (% , *n* = 10) | 38.5 | 50.0 |  |

\*t-values for continuous variables, $\chi^2$ for binary variables, adjusted residuals for categorical variables
\*\*p-values that maintained significance after Holm-Bonferroni correction are bolded

**Supplementary Table 8:** Neuroanatomic Regions of Interest in Participants without Hyperorality by Disease Stage.

|  | Mean Volume<br>[Standard Deviation]<br>(cm<sup>3</sup>) | Advanced<br>Stage<br>(cm<sup>3</sup>) | *p*-value |
| --- | --- | --- | --- |
| Right Lateral Orbitofrontal Cortex | 2802.3 [558.8] | -183.6 | 0.23 |
| Right Medial Orbitofrontal Cortex | 1677.0 [357.0] | -65.0 | 0.51 |
| Right Insula Cortex | 3888.5 [719.6] | 7.1 | 0.97 |
| Right Dorsal Striatum | 4117.3 [856.7] | -7.8 | 0.97 |
| Right Ventral Striatum | 243.0 [49.1] | 2.5 | 0.84 |
| Total Intracranial Volume | 1498520 [181708.6] |  |  |

\*generalized linear regression controlled for age at visit and total intracranial volume

**Supplementary Table 9:** Neuroanatomic Regions of Interest in Participants with Hyperorality by Disease Stage.

|  | Mean Volume<br>[Standard Deviation]<br>(cm<sup>3</sup>) | Advanced<br>Stage<br>(cm<sup>3</sup>) | *p*-<br>value** |
| --- | --- | --- | --- |
| Right Lateral Orbitofrontal Cortex | 2632.7 [655.8] | -327.6 | 0.03 |
| Right Medial Orbitofrontal Cortex | 1526.1 [388.5] | -183.5 | 0.05 |
| Right Insula Cortex | 3604.1 [826.8] | -342.0 | 0.09 |
| Right Dorsal Striatum | 3674.1 [1020.5] | -402.0 | 0.11 |
| Right Ventral Striatum | 222.6 [60.4] | -25.6 | 0.06 |
| Total Intracranial Volume | 1472910 [169273.4] |  |  |

\*generalized linear regression controlled for age at visit and total intracranial volume
\*\*p-values that maintained significance after Holm-Bonferroni correction are bolded

**Supplementary Table 10:** Neuroanatomic Changes and Hyperorality in Early and Advanced bvFTD.

|  |  | Mean Volume<br>[Standard<br>Deviation]<br>(cm<sup>3</sup>) | HYP+<br>coefficient<br>(cm<sup>3</sup>) | Hyp +<br>Coefficient 95%<br>CI | *p*-value** |
| --- | --- | --- | --- | --- | --- |
| CDR FTLD ≤1 | Right Lateral Orbitofrontal Cortex | 2872.1 [556.0] | -288.7 | -700.7 – 123.3 | 0.16 |
|  | Right Medial Orbitofrontal Cortex | 1698.6 [376.4] | -195.3 | -451.1 – 60.4 | 0.13 |
|  | Right Insula Cortex | 3885.8 [775.2] | -63.4 | -641.4 – 514.6 | 0.82 |
|  | Right Dorsal Striatum | 4126.7 [956.5] | -345.2 | -1010.9 – 320.4 | 0.30 |
|  | Right Ventral Striatum | 243.9 [55.1] | -17.6 | -54.6 – 19.4 | 0.34 |
|  | Total Intracranial Volume | 1491773 [171465.2] |  |  |  |
| CDR FTLD >1 | Right Medial Orbitofrontal Cortex | 2599.2 [628.40] | -98.0 | -500.0 – 303.9 | 0.63 |
|  | Right Lateral Orbitofrontal Cortex | 1530.5 [362.3] | -64.3 | -297.7 – 169.1 | 0.58 |
|  | Right Insula Cortex | 3642.3 [769.0] | -246.4 | -700.5 – 207.8 | 0.28 |
|  | Right Dorsal Striatum | 3757.3 [971.9] | -280.7 | -914.0 – 352.5 | 0.38 |
|  | Right Ventral Striatum | 225.1 [55.5] | -14.2 | -46.6 – 18.2 | 0.38 |
|  | Total Intracranial Volume | 1481916 [188277.8] |  |  |  |

\*generalized linear regression controlled for age at visit, sex, genetic status, and total intracranial volume
\*\**p*-values that maintained significance after Holm-Bonferroni correction are bolded

## Notes

### Author Declarations

Johns Hopkins School of Medicine Institutional Review Board approved this study under IRB00227492 on January 8th 2020.

